# Detection of asymptomatic *Leishmania* infection in Bangladesh by novel antigen and antibody diagnostic tools shows strong association with PKDL patients

**DOI:** 10.1101/2020.05.26.20113431

**Authors:** Sophie Owen, Faria Hossain, Prakash Ghosh, Rajashree Chowdhury, Md. Sakhawat Hossain, Christopher Jewell, Isra Cruz, Albert Picado, Dinesh Mondal, Emily R. Adams

## Abstract

**Background:** Asymptomatic *Leishmania* infections outnumber clinical infections on the Indian sub-continent (ISC) where disease reservoirs are anthroponotic. Diagnostics which detect active asymptomatic infection, which are suitable for monitoring and surveillance, may be of benefit to the visceral leishmaniasis (VL) elimination campaign on the ISC.

**Methodology/Principal Findings:** Quantitative polymerase chain reaction (qPCR), loop mediated isothermal amplification (LAMP), the direct agglutination test (DAT), and the *Leishmania* antigen ELISA were carried out on blood and urine samples collected from 720 household and neighbouring contacts of 276 VL and post kala-azar dermal leishmaniasis (PKDL) index cases, with no symptoms or history of VL and PKDL, in endemic regions of Bangladesh between September 2016 and March 2018. Of the 720 contacts of index cases, asymptomatic infection was detected in 69 (9.6%) participants by a combination of qPCR (1.0%), LAMP (2.1%), DAT (3.9%), and *Leishmania* antigen ELISA (3.3%). Only 1 (0.1%) participant was detected positive by all 4 diagnostic tests. Poor agreement between tests was calculated using Cohen’s kappa (κ) statistics, however the *Leishmania* antigen ELISA and DAT in combination capture all participants positive by more than one test. We find strong evidence for association between the index case being a PKDL case (OR 1.94, p = 0.009), specifically macular PKDL (OR 2.12, p = 0.004) and being positive for at least one of the four tests.

**Conclusions/Significance:** *Leishmania* antigen ELISA detects active asymptomatic infection, requires a non-invasive sample, and therefore may be of benefit for monitoring transmission and surveillance in an elimination setting in combination with serology. Development of an antigen detection test in RDT format would be of benefit to the elimination campaign.

**Author summary:** Infection with the parasite *Leishmania donovani* can lead to an asymptomatic infection with only around 5% of asymptomatics converting to visceral leishmaniasis the clinical manifestation of the infection. Serological assays detect anti-*Leishmania* antibodies and therefore cannot distinguish between past and active infection. Molecular assays and those which detect *Leishmania* antigens detect active infection. Since the signing of a memorandum of understanding in 2005, visceral leishmaniasis has been targeted for elimination in India, Nepal and Bangladesh. In an elimination setting such as Bangladesh, where disease reservoirs are anthroponotic, a relatively simple test such as the *Leishmania* antigen ELISA which requires a non-invasive urine sample, may be of benefit in combination with serology for surveillance and monitoring of *Leishmania* transmission. Development of an antigen test into a field compatible rapid diagnostic test would be of further benefit to the elimination campaign.

## Introduction

Infection with the parasite *Leishmania donovani (L. donovani)* usually manifests as asymptomatic infection with a small risk of progression to visceral leishmaniasis (VL), which in the absence of treatment is considered fatal [1]. Progression from asymptomatic infection to symptomatic disease was estimated to be between 5.6 and 15.2% in individuals with high anti-*Leishmania* antibody titers, as measured by the direct agglutination test (DAT), in India and Nepal [2], Globally, the ratio of asymptomatic to symptomatic VL varies [3]. In Bangladesh, the number of asymptomatic cases were found to outnumber symptomatic cases by 4:1 between 2002 and 2004 **[4]**.

Asymptomatic infection is of importance to VL endemic regions of the Indian sub-continent (ISC - India, Nepal, and Bangladesh) where the disease has been the target of an elimination campaign since 2005 [5,6]. Epidemiology of VL is cyclical and outbreaks occur approximately every 15 years on the ISC [7]. Asymptomatic carriers may represent a potential source of transmission in a region where parasite reservoirs are anthroponotic [8]. However, it is yet to be determined whether asymptomatically infected humans are infective to sand flies. A study in a small number of asymptomatically infected dogs showed that parasites were transmittable to sand flies [9], however no human data has yet been recorded. Sixteen (8.2%) asymptomatic individuals who converted to VL within 2 years in a study in Bangladesh were found to have significantly higher anti-rK39 antibody titers compared to their counterparts who did not progress [10].

The rK39 enzyme-linked immunosorbent assay (ELISA), rK39 rapid diagnostic test (RDT) and the DAT measure the presence of anti*-Leishmania* antibodies [11-14], These antibodies have been found to persist for months or years after infection, with patients in the VL endemic region of Muzaffarpur, India found positive by rK39 RDT (39.0%) and DAT (53.0%) ≥ 15 years post treatment [15]. Therefore, a clinical history is required to determine if a positive result is due to active or previous infection, or a previous asymptomatic infection that will not progress to disease. Tests which detect active infection, such as quantitative real-time polymerase chain reaction (qPCR), loop mediated isothermal amplification (LAMP) or *Leishmania* antigen ELISA could be used as tools to monitor active asymptomatic infection, and quickly identify areas with increasing active transmission.

Highly sensitive qPCR was shown to be an effective technique for diagnosis of VL and monitoring of treatment response, and could be of value in an elimination setting [16]. The *Leishmania* antigen ELISA (Clin-Tech, Guilford, UK) detects low molecular weight *Leishmania* carbohydrates excreted in the urine and therefore detects active infection and uses a non-invasive sample type. A study found sensitivity to range from 77-87% across endemic countries [17]. Loop-mediated isothermal amplification (LAMP) enables the robust, fast, simple and highly specific amplification of nucleic acids and does not require a thermocycler or cold-chain; Loopamp™ *Leishmania* Detection Kit (Eiken Chemical Co., Japan) targets both the 18S rDNA and kinetoplast DNA (kDNA) and was previously demonstrated to have a sensitivity of 92% in VL suspect patients in Ethiopia [18]. Similarly high sensitivities of 98% and 100% were seen in a study in Sudan with the Loopamp™ *Leishmania* Detection Kit (Eiken Chemical Co., Japan) when DNA was extracted from peripheral blood using boil and spin and QIAamp DNA mini kits (Qiagen, Hilden, Germany) respectively [19].

To determine the utility of kDNA qPCR, Eiken LAMP, and the *Leishmania* antigen ELISA for detection of active asymptomatic infection in an elimination setting, we tested samples collected from household or neighbouring contacts of index cases from endemic regions of Bangladesh. This paper presents estimates of disease state, sensitivity, and specificity of the diagnostic tests using latent class analysis (LCA) and risk factors for asymptomatic infection.

## Methods

### Study ethics

This study was approved by the Ethical Review Committee (ERC) of the ICDDR,B (PR-14093). Adult participants provided written informed consent, and in the case of any participants under 18 years of age, a parent or guardian provided informed consent.

### Asymptomatic visceral leishmaniasis clinical samples

Blood and urine samples from 720 clinically healthy household and neighbouring contacts in adjacent households of 276 VL or PKDL index cases (between 1 and 8 contacts per index case), aged 5 to 60 years, with no symptoms or history of VL and PKDL, were collected between September 2016 and March 2018. The study was conducted in the VL endemic districts of Mymensingh, Gazipur, Tangail, Narail, Jamalpur, Pabna, and Brahmanbaria in Bangladesh. Symptoms considered included presence of fever, rash, loss of appetite, weight loss, lymph node enlargement, abdominal enlargement and pain. Demographic and clinical parameters were recorded. Samples were transported to Dhaka using a cold chain for processing and laboratory analysis using DAT and LAMP. Urine and DNA samples for *Leishmania* antigen ELISA and qPCR respectively were transported on ice to the UK from Bangladesh and stored at -20°C until testing.

### DNA extraction

DNA was extracted in three different ways: 1. DNA was extracted from 100μl whole blood and eluted in 200μl buffer using DNeasy blood and tissue DNA extraction kits (Qiagen, Hilden, Germany) as per the manufacturer’s instructions. 2. Boil and spin extractions were carried out by pre-treating whole blood samples with SDS. Briefly, 10% SDS solution was mixed with blood to a final concentration of 5% and stored at -20°C. Once defrosted, samples were inverted 10 times and allowed to stand at room temperature for 10 minutes. Samples were further inverted and 400μl of distilled water were added before incubation at 90°C for 10 minutes. Tubes were then centrifuged at maximum speed for 3 min and the supernatants stored for testing. 3. DNA was extracted from dried blood spots (DBS). Whole blood was air-dried onto Whatman filter paper (GE Healthcare Life Sciences, Buckinghamshire, UK) for 30 min at room temperature and stored in individual bags. Discs of 7mm were punched out of the paper and added to an Eppendorf with 50μl of double-distilled water. Tubes were incubated at 90°C for 10 minutes followed by centrifugation for 3 minutes at maximum speed. Supernatants were stored at -20°C for testing.

### qPCR

Real time PCR (qPCR) was performed on DNA extracted from whole blood using Qiagen DNeasy kits (Qiagen, Germany) [18]. A 1.25 μL volume of DNA was added to 11.25 μL amplification mixture containing 2.5 μL QuantiFast mastermix (Qiagen, Germany), 0.4 μM kDNA forward primer, 0.4 μM kDNA reverse primer, and 0.2 μM kDNA FAM-probe. Amplification was performed on a Qiagen Rotor-Gene Q system with following reaction conditions: 5 min at 95°C, followed by 40 cycles of 15 seconds at 95°C and 30 seconds at 60°C. Data was analysed using the Rotor-Gene Q series software (Qiagen, Germany). Standard curve analysis was performed using *Leishmania donovani* DNA (positive control) and data was used to set a qPCR threshold. Samples with cycle threshold (Ct) <34 were considered positive.

### Loop mediated isothermal amplification (LAMP)

LAMP was run on DNA extracted from whole blood using the DNeasy blood and tissue DNA extraction kits (Qiagen, Germany), boil and spin extraction, and extraction from dried blood spots as described above. Loopamp™ *Leishmania* Detection kits (Eiken Chemical Co., Ltd, Tokyo, Japan) were used. Samples to be tested were made up to total volume of 30μl by adding 3μl DNA sample to 27μl of water. The lids of the tubes were then closed, and the sample mixed with the master mix contained in the tube cap by inverting the tubes and leaving to stand for 2 minutes cap-side down. The tubes were inverted 5 times, spun down and incubated at 65°C for 40 minutes, then 80°C for 5 minutes. Results were visualized under blue LED light illumination, using the Fluorescence Visual Check Unit of the HumaLoop M incubator (HUMAN, Wiesbaden, Germany). Results were read by two technicians blinded to each other. A third technician was used in the event of disagreement and the majority used.

### Direct agglutination test (DAT)

The DAT was carried out in Bangladesh and performed as previously described [20].

Following a dilution of sera 1:100, the samples were further diluted in eight two-fold serial dilutions. Where samples did not react in the first dilution, the end titer was read as <1:200. Where samples still reacted at the final dilution, the end titer was read as >1:25,600. The threshold for a positive DAT result was set at ≥1:1600 as previously used by Hasker *et al*. for detection of asymptomatic infection [20].

### *Leishmania* antigen ELISA

The *Leishmania* antigen ELISA (Clin-Tech., Guilford, UK) was performed on urine samples as per the manufacturer’s instruction. Briefly, samples were diluted 1 in 20 with assay diluent. 100μl of antigen calibrators and diluted samples were added to a pre-coated 96-well plate and incubated at 37°C for 30 minutes. Following 4 washes, 100μl of working strength tracer was added to the wells and incubated at 37°C for 30 minutes. Following a further 4 washes, 100μl of TMB substrate was added to each well and incubated uncovered between 18° and 25°C for 30 minutes. 100μl of stop solution was then added to each well. A standard curve was included on each plate. The optical densities (OD) were read at 450nm and blanked on air or with the 620nm reading within 30 minutes of addition of stop solution. Four parameter curve fitting software was used to calculate the concentration (UAU/ml) of each sample. IBM SPSS Statistics 24 was used to generate receiver-operating characteristic (ROC) curves using 720 asymptomatic cases and 80 VL cases to determine the threshold in UAU/ml that gave a sensitivity of 98.8% and a specificity of 96.7%. The area under the curve (AUC) was calculated.

### Statistical analysis

Data were analysed in R Studio Version 1.1.456. Discrete variables were summarised as counts and percentages. Continuous variables were summarised as the median and interquartile range (IQR). The software package ‘Venny’ was used to create Venn diagrams for comparison of diagnostic tests [21].

Percentage agreement between diagnostic tests and Cohen’s kappa (κ) statistics with p-values to measure agreement between diagnostic tests were calculated with the irr package version 0.84.1 in R. Logistic regression was used to regress asymptomatic *L. donovani* infection (defined as positive for at least one of the four tests) outcome variable onto potential risk factor variables identified in the literature. Latent class analysis was used to estimate diagnostic accuracy and prevalence [22], Test results were assumed to be conditionally dependent, with Bayesian prior distributions on sensitivity, specificity and prevalence set using Betabuster 1.0 (https://betabuster.software.informer.com/). The analysis was implemented in R Studio Version 1.1.456 using the ‘lcaR’ model written by Jonathan Marshall (version 2bc8ca6, 13^th^ November 2015) [23].

## Results

### Study population

A total of 720 individuals were sampled, of those 280 (38.9%) were male with a median age of 27 (IQR = 25). Most common occupations were students (34.4%) and housewives (41.9%). A total of 505 (70.1%) contacts lived within the household of an index case and 215 (29.9%) lived within a neighbouring household.

A total of 69 individuals were positive for at least one diagnostic test. Of those, 31 (44.9%) were male with a median age of 30 (IQR = 25). Most common occupations within the 69 individuals were students (33.3%) and housewives (37.7%). 50 (72.5%) lived within the household of an index case. The 69 asymptomatic cases were spread across 59 (21.4%) of the 276 index cases. Of those 69 index cases, median percentage positivity of the contacts was 33.3% (IQR = 25).

The 720 contacts were associated with VL cases (66.1%) - made up of new VL cases (90.1%), relapsed VL (9.5%), VL treatment failure (0.4%) - or PKDL cases (33.9%) (Table 1). Of the 242 PKDL index cases with known rash type, 230 (95.0%) presented with macular rash, 4 (1.7%) with macular and papular rash, 6 (2.5%) with nodular and macular rash, and 2 (0.8%) with macular, nodular and papular rash (Table 1). The 69 asymptomatic cases were associated with new VL cases (49.3%) or PKDL cases (50.7%), with the majority of such PKDL cases presenting with macular rash (94.3%) (Table 1).

**Table 1.**
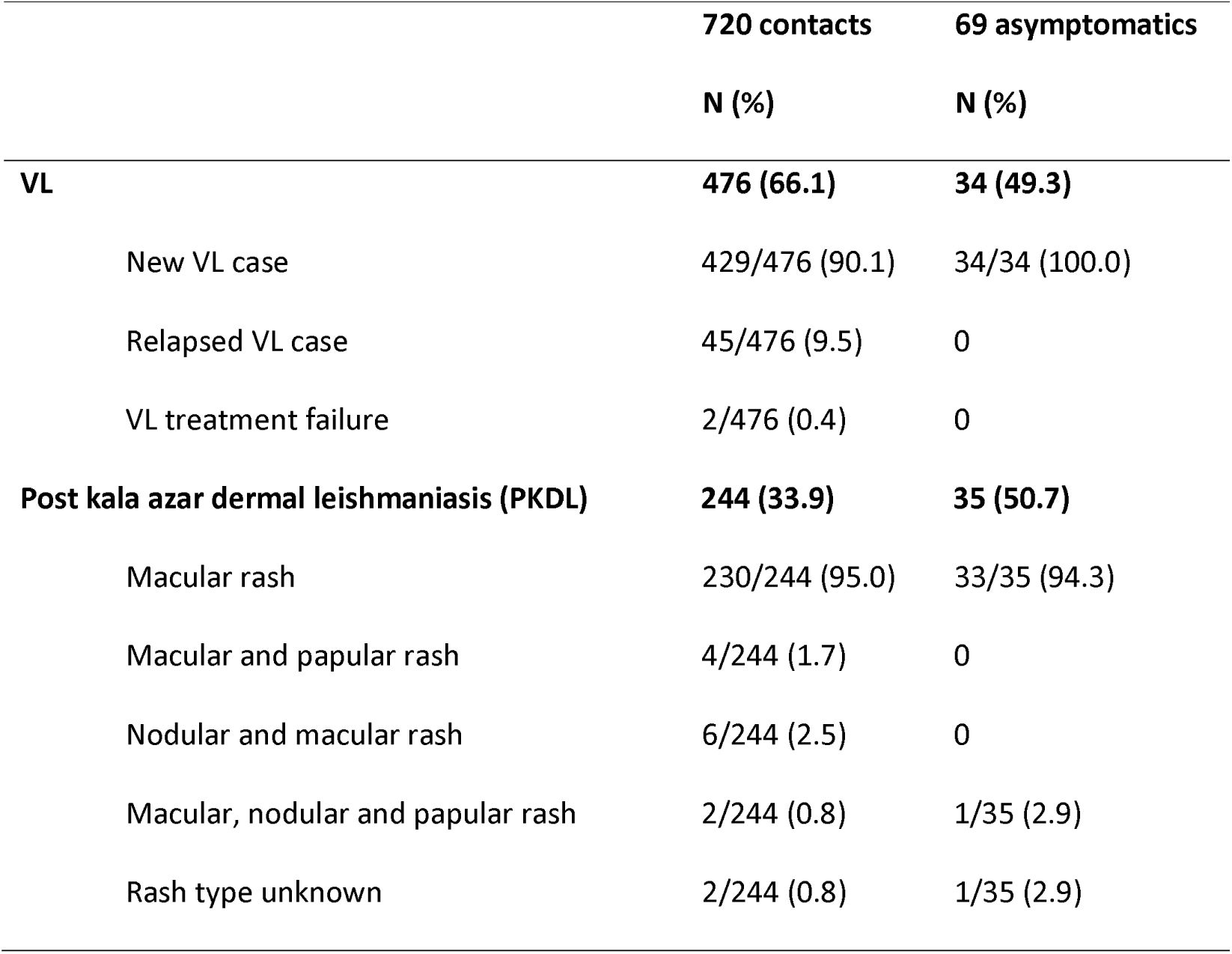
Index cases were classified as new VL cases, relapsed VL cases, VL treatment failure or PKDL. Of the 720 contacts, 476 (66.1%) were associated with VL cases and 244 (33.9%) were associated with PKDL cases. Of the 69 participants positive for at least one test, 34 (49.3%) were VL cases and 35 (50.7%) were PKDL cases.

### Estimates of asymptomatic infection in contacts of index cases using tests to detect active infection

Of the 720 participants screened, 69 (9.6%) were positive by at least one test. Of the 720 asymptomatic DNA samples screened, 7 (1.0%) were positive by kDNA qPCR, with a mean Ct value of 31.9 (range 26.7 - 33.9). Urine samples were screened with the *Leishmania* antigen ELISA, of which 24 (3.3%) were found to be positive. Samples screened by DAT were considered positive at a titer of ≥1:1600. A total of 28 (3.9%) samples were found to be DAT positive, 11 (39.3%) of which had a titer ≥1:12,800. LAMP detected 6 (0.8%), 8 (1.1%), and 3 (0.4%) asymptomatic infections when DNA was extracted using Qiagen kits, boil and spin, and from DBS, respectively. For the purposes of further analysis, a participant with a positive LAMP result from any one of the three extraction techniques was considered LAMP positive, of which there were 15 (2.1%).

*Leishmania* antigen ELISA and the DAT identified the highest proportion of positive subjects. Only 1 (0.1%) subject was identified as positive by all 4 diagnostic methods, 2 (0.3%) were identified by 2 diagnostic methods, and 66 (9.2%) were identified by 1 diagnostic method only. In the 69 asymptomatic participants, 26 (37.7%) were positive by DAT only and 6 (8.7%) were positive by qPCR only. Of the 24 (34.8%) participants positive by ELISA, 3 (4.3%) were positive by at least one other test (Fig 1). Generally, poor agreement was found between tests. However, antigen and molecular tests showed better agreement in combination compared to the same tests in combination with serology (Table 2). In combination, the DAT and *Leishmania* antigen ELISA capture all participants positive by more than 1 of the 4 tests.

**Fig 1.**
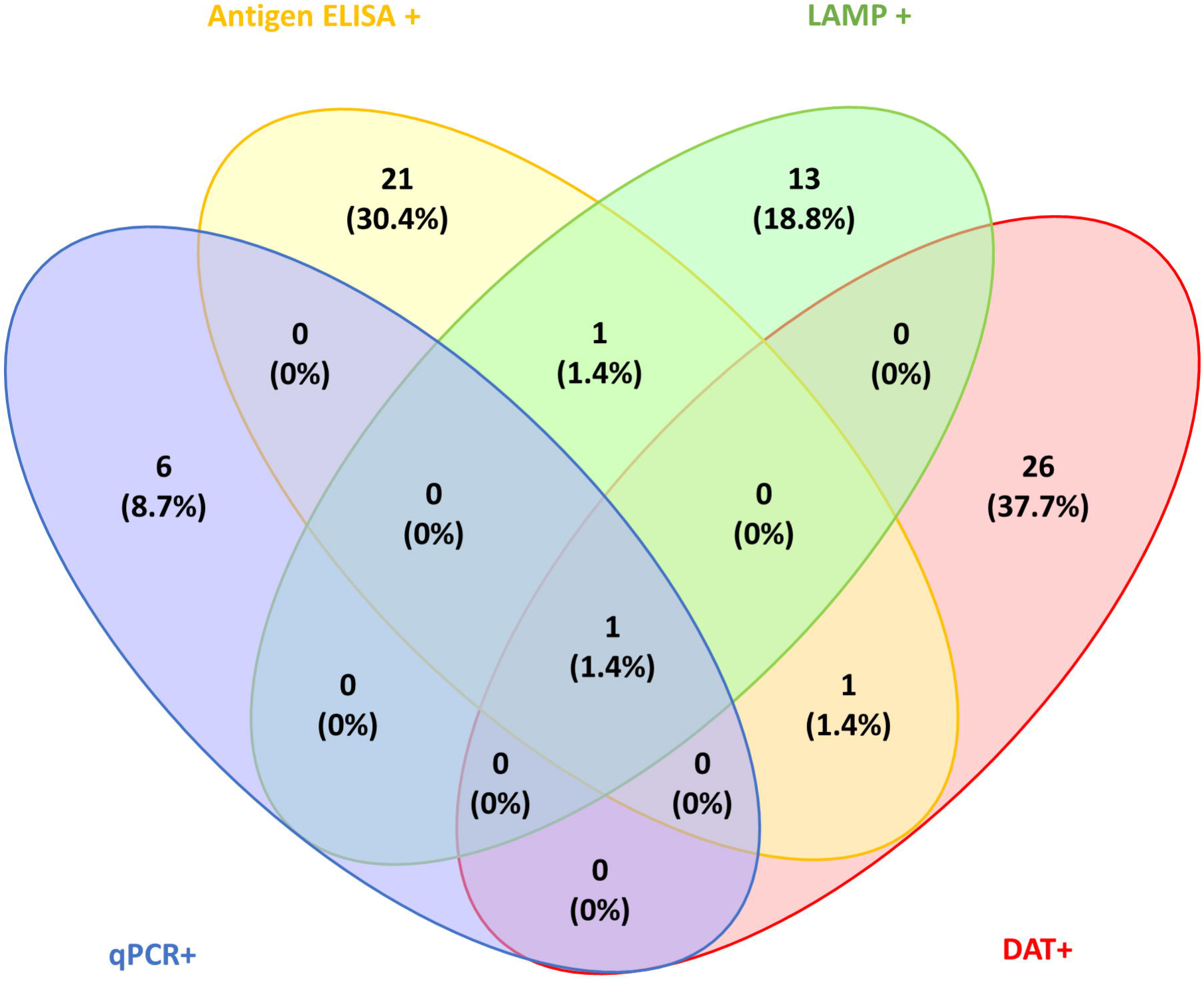
Asymptomatic infection was detected in 69 (9.6%) contacts by a combination of four diagnostic tests. DAT was positive in 28 (40.6%) participants, 26 (37.7%) of whom were positive for DAT alone, 11/28 (39.3%) of whom had a titer greater than 1:12,800. QPCR was positive in 7 (10.1%) participants, 6 (8.7%) of whom were positive for qPCR alone. LAMP was positive in 15 (21.7%) participants, 13 (18.8%) of whom were positive for LAMP alone. *Leishmania* antigen ELISA was positive in 24 (34.8%) participants, of whom 21 (30.4%) were positive for ELISA alone and 3 (4.3%) were positive by ELISA and at least 1 other test.

**Table 2.**
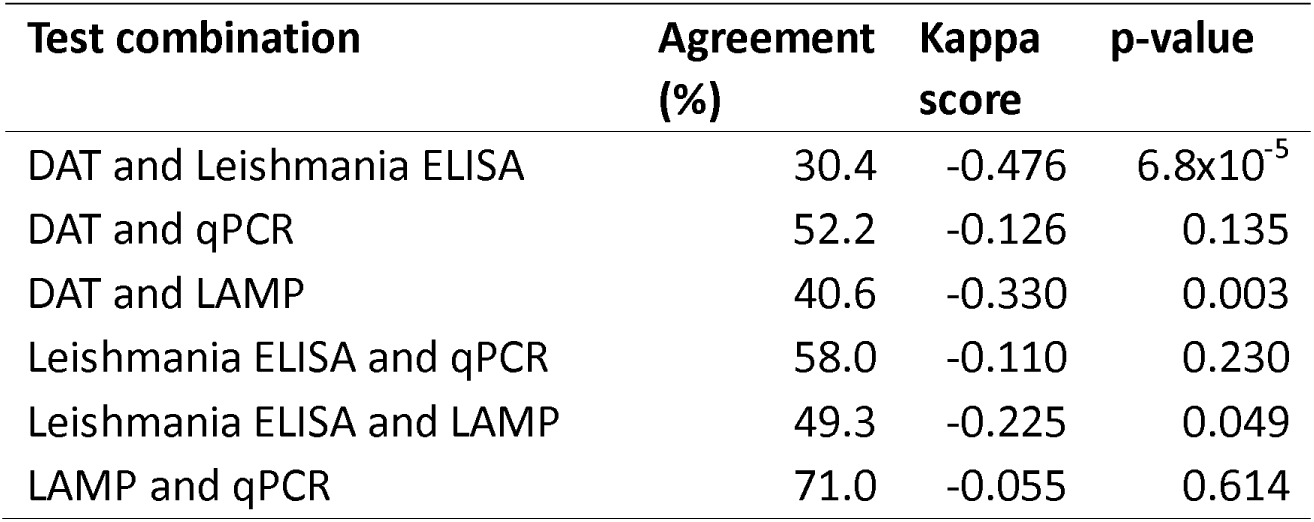
Kappa scores and agreement for 4 diagnostic tests in 69 asymptomatic participants.

| Test combination | Agreement (%) | Kappa score | p-value |
| --- | --- | --- | --- |
| DAT and *Leishmania* ELISA | 30.4 | -0.476 | $6.8 \times 10^{-5}$ |
| DAT and qPCR | 52.2 | -0.126 | 0.135 |
| DAT and LAMP | 40.6 | -0.330 | 0.003 |
| *Leishmania* ELISA and qPCR | 58.0 | -0.110 | 0.230 |
| *Leishmania* ELISA and LAMP | 49.3 | -0.225 | 0.049 |
| LAMP and qPCR | 71.0 | -0.055 | 0.614 |

### Risk factors for asymptomatic VL

Logistic regression was used to confirm risk factors associated with being positive for at least one of the four diagnostic tests. Age, gender, occupation, and living within the index household compared to neighbouring household were not found to be associated with asymptomatic infection. The index case being a PKDL case (OR 1.94, p = 0.009), specifically macular PKDL (OR 2.12, p = 0.004) were found to be significantly associated with being positive by at least one of the four tests.

### Latent class analysis to estimate disease state and diagnostic accuracy in the absence of a gold standard

In the absence of a single reference standard or a composite reference standard, latent class analysis (LCA) was used to estimate disease state. LCA estimated qPCR, LAMP, DAT, and *Leishmania* antigen ELISA to have sensitivities (2.5 - 97.5 percentiles) of 85.6% (55.1 - 99.5), 99.8% (99.2 - 99.9), 97.5% (90.5 - 99.9), and 98.9% (96.2 - 99.9) and specificities of 96.1% (94.7 - 97.5), 96.7% (95.3 - 97.8), 99.0% (98.1 - 99.6), and 97.9% (96.7 - 98.9), respectively. Prevalence of *L. donovani* asymptomatic infection in VL and PKDL contacts in Bangladesh was estimated to be 0.3% (0.03 - 0.7).

## Discussion

In this study, we measured the prevalence of active asymptomatic VL infection in household or neighbouring contacts of VL and PKDL index cases by qPCR, *Leishmania* antigen ELISA, and LAMP in endemic regions of Bangladesh. PKDL cases are a potential reservoir of *Leishmania* infection with experimental infectivity to sand flies estimated to be between 32 and 53% [24], Here, we demonstrate a risk factor for asymptomatic infection is living close to a PKDL case, specifically macular PKDL. This follows the launch of the World Health Organisation’s road map for neglected tropical diseases 2021-2030, which identifies early detection through methods such as active case detection and development of treatments and diagnostics for both VL and PKDL, as critical actions for the elimination of VL as a public health concern [25]. Our data and the road map highlight the importance of diagnosis of PKDL cases and their follow-up, in recognition of their potential role in transmission.

Previous studies have identified risk factors for VL broadly linked to poverty, such as mud walls, with sleeping off the floor found to reduce the risk [26]. Proximity to a previous VL case was identified as a risk factor for VL in Bangladesh [27], No difference based on sex, occupation, or income among others, was seen in an analysis of risk factors in the same study [27], Age trends associated with VL infection were found to vary between studies, however prevalence of sero- positivity was generally found to increase with age [28]. We acknowledge that sample size may have limited our analysis of risk factors. A limitation of the study includes the lack of follow-up data, therefore the accuracy of the tests as predictors of progression to clinical disease is unknown.

Both the DAT and *Leishmania* antigen ELISA capture all samples which are positive by more than one test and both utilise sample types that have a relatively non-invasive sample collection, which can be transported back to a central laboratory for testing. The specificity of all diagnostics falls below 100% for identification of *L. donovani* asymptomatic infection, therefore we expect some false positives on a cohort of this size; therefore, we have looked for overlap in tests which were positive.

The DAT detected the highest proportion of positive individuals. The DAT detects anti-*Leishmania* antibodies that could be circulating from a previously cleared asymptomatic infection. However, a recent study found that DAT titers could be a useful tool to monitor transmission in an elimination setting during repeat surveys [14]. Where qPCR requires more laboratory infrastructure, the *Leishmania* antigen ELISA and LAMP are relatively simple techniques suitable for use in resource poor settings. Furthermore, *Leishmania* antigen ELISA requires a non-invasive urine sample which may aid in screening of high numbers of asymptomatic contacts. Since living with or close to a macular PKDL patient is a risk-factor for asymptomatic infection, we propose the follow-up of contacts with PKDL patients as an operational priority. Development of an antigen detection test in RDT format would be of benefit to identify those contacts.

## Data Availability

Data is available upon request.

## Acknowledgements

We would like to thank the field teams and the patients. Funding was received for this study from the German Federal Ministry of Education and Research (BMBF) through the KfW Entwicklungs bank, MRC-DTP (MR/N013514/1), and Wellcome Seed fund (108080/Z/15/Z). FIND is grateful to its donors, public and private, who have helped bring innovative new diagnostics for diseases of poverty. A full list of FIND’s donors can be found at: https://www.finddx.org/donors/.

## Notes

### Competing Interest Statement

The authors have declared no competing interest.

### Author Declarations

This study was approved by the Ethical Review Committee (ERC) of the ICDDR,B (PR-14093).

